# Harmonising UK primary care prescription records for research: A case study in the UK Biobank

**DOI:** 10.64898/2026.04.21.26351274

**Authors:** Cai R Ytsma, Ana Torralbo, Natalie K Fitzpatrick, Maik Pietzner, Ioannis Louloudis, Sabrina Ansarey, Daniela Nguyen, Spiros Denaxas

## Abstract

**Objective:** The aim of this study was to develop and validate an automated, scalable framework to harmonise fragmented UK primary care prescription records into a research-ready dataset by mapping four diverse medical ontologies to a unified, historically comprehensive reference standard.

**Materials and Methods:** We used raw prescription records for consented participants in the UK Biobank, in which participants are uniquely characterized by multiple data modalities. Primary care data were preprocessed by selecting one drug code if multiple were recorded, cleaning codes to match reference presentations, expanding code granularity based on drug descriptions, and updating outdated codes to a single reference version. Harmonisation entailed mapping British National Formulary (BNF) and Read2 codes to dm+d, the universal NHS standard vocabulary for uniquely identifying and prescribing medicines. Harmonised dm+d records were then homogenised to a single concept granularity, the Virtual Medicinal Product (VMP). We validated our methods by creating medication profiles mapping contemporary drug prescribing patterns in 312 physical and mental health conditions.

**Results:** We preprocessed 57,659,844 records (100%) from 221,868 participants (100%). Of those, 48,950 records were dropped due to lack of drug code. 7,357,572 records (13%) used multiple ontologies. Most (76%) records were encoded in BNF and most had the code granularity expanded via the drug description (N=28,034,282; 49%). 41,244,315 records (72%) were harmonised to dm+d and 99.98% of these were converted to VMP as a homogeneous dataset. Across 312 diseases, we identified 23,352 disease-drug associations with 237 medications (represented as BNF subparagraphs) that survived statistical correction of which most resembled drug - indication pairs.

**Conclusion:** Our methodology converts highly fragmented and raw prescription records with inconsistent data quality into a streamlined, enriched dataset at a single reference, version, and granularity of information. Harmonised prescription records can be easily utilised by researchers to perform large-scale analyses in research.

## BACKGROUND AND SIGNIFICANCE

Electronic health records (EHR), data generated as part of clinical care interactions, contain a wealth of information for researchers including detailed medication prescription data. Medication information can help researchers to identify cohorts of people who are prescribed certain drugs, understand trends in prescribing behavior related to shifts in healthcare system policy and reward schemes, or analyse the cost burden of drugs prescribed in primary or secondary care. In general, prescription data enable scientists to uncover meaningful insights into the health and behaviour of the nation, in particular if linked to expensive research data such as whole-genome sequencing [1]. Additionally, accurate medication prescribing profiles for diseases are a valuable resource that can inform policy makers and accelerate drug discovery by identifying potential drug repurposing opportunities. Medication prescription information however is often fragmented and not stored in a single unified source or format.

In the United Kingdom, primary care prescriptions are recorded using three different medical ontologies and require a significant amount of pre-processing, cleaning, and harmonisation to become research-ready. In many cases, local medical ontologies are not widely used standards which further complicates international analyses spanning multiple biobanks. Finally, no single source of pragmatic medication prescribing patterns exists for physical and mental health conditions diagnosed in UK primary care as these are often analysed in a single or small set of diseases.

In this study, we provide a reproducible methodology for harmonising primary care prescriptions from England, Scotland, and Wales in UK Biobank participants by mapping recorded medications to an international standard and a common structure and format. Furthermore, for 312 physical and mental health conditions we provide an open-access resource of contemporary medication prescribing profiles derived from primary care EHR data.

## MATERIALS AND METHODS

### data sources

We used data from the UK Biobank (UKB), a longitudinal study of 500,000 middle aged adults tracking the health of people in the UK as they age. Volunteers aged 40-69 attended assessment centres in England, Scotland, and Wales between 2006-2010 to fill out self-reported questionnaire data, undergo baseline assessment of physiological measurements, and consent to sharing of EHR for study follow-up. Of the entire UKB study, primary care prescription records are available for approximately 220,000 [2] individuals with the remaining data for the entire cohort estimated to be released in 2026. The UKB is one of the most widely used open access research datasets in the world with over 22,000 approved researchers in more than 60 countries.

#### Study population

The study population consisted of UKB participants with linkage to primary care data that had at least one valid primary care registration period (defined as a valid date of practice registration) in a primary care practice and at least one prescription event recorded, spanning from 26 July 1945 to 25 August 2019. Most records commenced in the late 1980s with only 279 events recorded between 26 July 1945 and 1 January 1986. Prescription records are recorded in four primary care data provider EHR systems which are described below.

#### Prescription data providers

We included data from four UKB primary care data providers: two from England (TPP and Vision), one from Scotland and one from Wales. Providers each use one or two out of three ontologies to encode their prescriptions (**Table 1**). UKB receives the data via various suppliers (e.g., Albasoft for Scottish data, SAIL Databank for Welsh data) and performs minimal pre-processing to the raw prescription records, in order to retain the received information as closely as possible [3]. This included replacing event dates with dummy dates when a) the date precedes the participant date of birth (replaced with 01/01/1901), b) the date matches the participant date of birth (02/02/1902), c) the date is within the year of birth of the participant (03/03/1903), or d) the date is in the future (07/07/2037). These steps were performed by UKB before distributing data to researchers.

**Table 1.** Summary of raw UK Biobank primary care prescription records stratified by ontology and data provider. *One entry may record drugs in multiple ontologies. Raw Welsh records using Read2 did not contain any drug names. See Supplementary Figure S1 for overlap.

| Data provider |  | All records | dm+d* | BNF* | Read2* |
| --- | --- | --- | --- | --- | --- |
| All | N records | 57,659,844 | 6,343,919 | 43,733,704 | 14,890,845 |
|  | N people | 221,868 | 18,271 | 184,572 | 51,964 |
|  | N unique codes | - | 13,318 | 5,201 | 19,522 |
| 1 (England Vision) | N records | 6,343,919 | 6,343,919 | - | 6,220,687 |
|  | N people | 18,271 | 18,271 | - | 18,266 |
|  | N unique codes | - | 13,318 | - | 10,268 |
|  | N unique drug names | - | 41,794 | - | 34,094 |
| 2 (Scotland) | N records | 4,298,107 | - | 4,249,157 | 1,136,887 |
|  | N people | 24,579 | - | 24,579 | 13,253 |
|  | N unique codes | - | - | 4,566 | 4,514 |
|  | N unique drug names | - | - | 21,415 | 5,580 |
| 3 (England TPP) | N records | 39,484,547 | - | 39,484,547 | - |
|  | N people | 160,018 | - | 160,018 | - |
|  | N unique codes | - | - | 635 | - |
|  | N unique drug names | - | - | 23,344 |  |
| **4 (Wales)** | N records | 7,533,271 | - | - | 7,533,271 |
|  | N people | 20,483 | - | - | 20,483 |
|  | N unique codes | - | - | - | 9,579 |
|  | N unique drug names | - | - | - | - |

#### Prescription ontologies

There are three ontologies represented in the UKB primary care prescription data: Read2, dm+d and BNF. Information about the ontologies, including documentation, lookup, and mapping files, are published by the NHS Business Services Authority (NHSBSA) [4]. A breakdown of ontologies recorded per data provider and the overlap of ontologies within the raw entries are shown in Table 1 and **Supplementary Figure S1**, respectively.

#### Read version 2 (Read2)

Read codes were first developed as a clinical ontology in the early 1980s and were widely used in UK primary care from 1999-2018. Read version 2 (Read2) codes were predominantly used for diagnoses and prescriptions until April 2016 [5]. At this time, the National Health Service (NHS) migrated to Clinical Terms Version 3 (CTV3) until its last release in April 2018, when SNOMED CT (which includes all CTV3 and Read version 2 codes by definition) became the official standard for recording information in the NHS [5]. Read2 codes are formatted as five case-sensitive characters: for example, ‘Paracetamol 500mg tablets’ are encoded as ‘di21.’. Read2 codes are used by all UKB data providers except England TPP. The static lookup file for Read2 codes is accessible from NHSBSA [6].

#### SNOMED-CT Dictionary of medicines and devices (dm+d)

In 2017, the NHS chose the SNOMED CT-based dictionary of medicines and devices (dm+d) as the standard for prescriptions in response to the “lack of standardisation in the UK in describing medicines…and in linking knowledge…to these descriptions” [7]. Similar to SNOMED CT, concepts in dm+d are represented by a unique numeric SNOMED CT concept identifier (SCTID).

There are five types of dm+d concepts as described in **Supplementary Table S1** and visualised in the NHSBSA data model [8]: Virtual Therapeutic Moiety (VTM); Virtual Medicinal Product (VMP); Virtual Medicinal Product Pack (VMPP); Actual Medicinal Product (AMP), and; Actual Medicinal Product Pack (AMPP). In the 2019 UKB primary care data release, Vision (England) is the only primary care data provider to encode prescriptions in dm+d, either at the VMP or AMP level. For example, the VMP code for ‘Paracetamol 500mg tablets’ is 42109611000001109; it has at least one associated AMP code, like 17953211000001103 which represents ‘Paracetamol 500mg tablets (Phoenix Healthcare Distribution Ltd)’.

##### Creating a historically comprehensive dm+d lookup

VMP (and VTM) codes can potentially change over time, e.g., when a medication is discontinued or a supplier is renamed. These changes are accounted for at the time of prescribing but historical records are not retrospectively updated. The NHS TRUD regularly releases updated and revised versions of ontologies used by the NHS, including dm+d (SNOMED CT) [4]. To address this challenge, we created a comprehensive dm+d lookup file that included all terminology releases spanning the study period which ended 25 August 2019: 2014 version 1.0.0 (the first dm+d release) to 2024 version 7.2.0 (released at the time of writing).

To develop and streamline a consolidated dm+d lookup file that contains all historical release information we obtained all original dm+d release files from NHS TRUD and deduplicated the lookup concept information while retaining the dm+d version where the concept was first and last mentioned. When dm+d concept identifiers change, records in release files that reference the code are replaced. With this information, we mapped identical drug concepts to their most recent version (at the time of work, 2024 version 7.2); the ‘reference’ code for the concept. These reference codes were used during harmonisation to align all historical concepts to a single version. A worked example of selecting a single reference concept for dm+d codes of the same concept using this consolidated lookup is visualised in **Supplementary Figure S2** with a detailed example in **Supplementary Table S2**. These reference concepts were also used when mapping BNF and Read2 to dm+d.

#### British National Formulary (BNF)

The British National Formulary (BNF) is an authoritative reference that provides evidence-based guidance on prescribing, pharmacology, and the safe use of medicines across the UK. BNF is accredited by the National Institute for Health and Care Excellence (NICE) and published by *The BMJ* [9]. It was used as the basis for the NHS Prescription Services, which calculate and report costs of prescribed and dispensed medications across England [10], until the adoption of dm+d in 2019 [11]. BNF code information is available from the NHS Business Services Authority (NHSBSA) Open DataPortal [12]. Within the UKB primary care prescription data, records from the Scottish and English TPP providers are encoded using BNF codes.

BNF codes are hierarchically organised and expansive, with increasing characters providing increasing detail about the drug (like usage, chemical substance, or presentation). For example, the concept ‘Paracetamol 500mg tablets’ is coded as 0407010H0AAAMAM: the sequence of characters describes the BNF chapter (04: Central Nervous System), section (0407: Analgesics), paragraph (040701: Non-opioid analgesics and compound preparations), chemical substance (0407010H0: Paracetamol), with final characters specifying the presentation of the drug (500mg tablets).

To map BNF codes to SNOMED CT dm+d codes, we extracted mappings from published mapping data [13] between BNF codes and all relevant dm+d codes. For example, a BNF code at a level of information equivalent to a dm+d VMP code is also mapped broadly to VMPP codes based on the VMP. For example, a BNF code for Verapamil 160mg tablets is mapped to the VMP code of the same concept, as well as to three VMPP codes that specify the number of tablets. For our study, we desired a simple 1:1 mapping of BNF to the most appropriate level of information in dm+d. Therefore, we deduplicated the published file by selecting the mapped dm+d code with the most equivalent concept (see **Supplementary Figure S3**). Finally, for a small number of remaining unmapped codes we expanded the map file by manually adding equivalent codes as identified by exactly matched descriptions. We then converted the mapped dm+d codes to ‘reference’ dm+d codes as described previously.

### preprocessing and harmonisation

For each prescription event, our framework (**Figure 1**) implements a two-step approach consisting of a data preprocessing stage (Figure 1A), and a harmonisation stage (Figure 1B). The preprocessing step cleaned raw ontology codes by selecting a single ontology code and aligning it with lookup files where necessary, while the harmonisation stage mapped the raw codes to dm+d codes. Descriptions and examples of each of the possible harmonisation scenarios are shown in **Figure 2**. We retained fields with event information such as participant identifier, issue date, and data provider.

**Figure 1.**
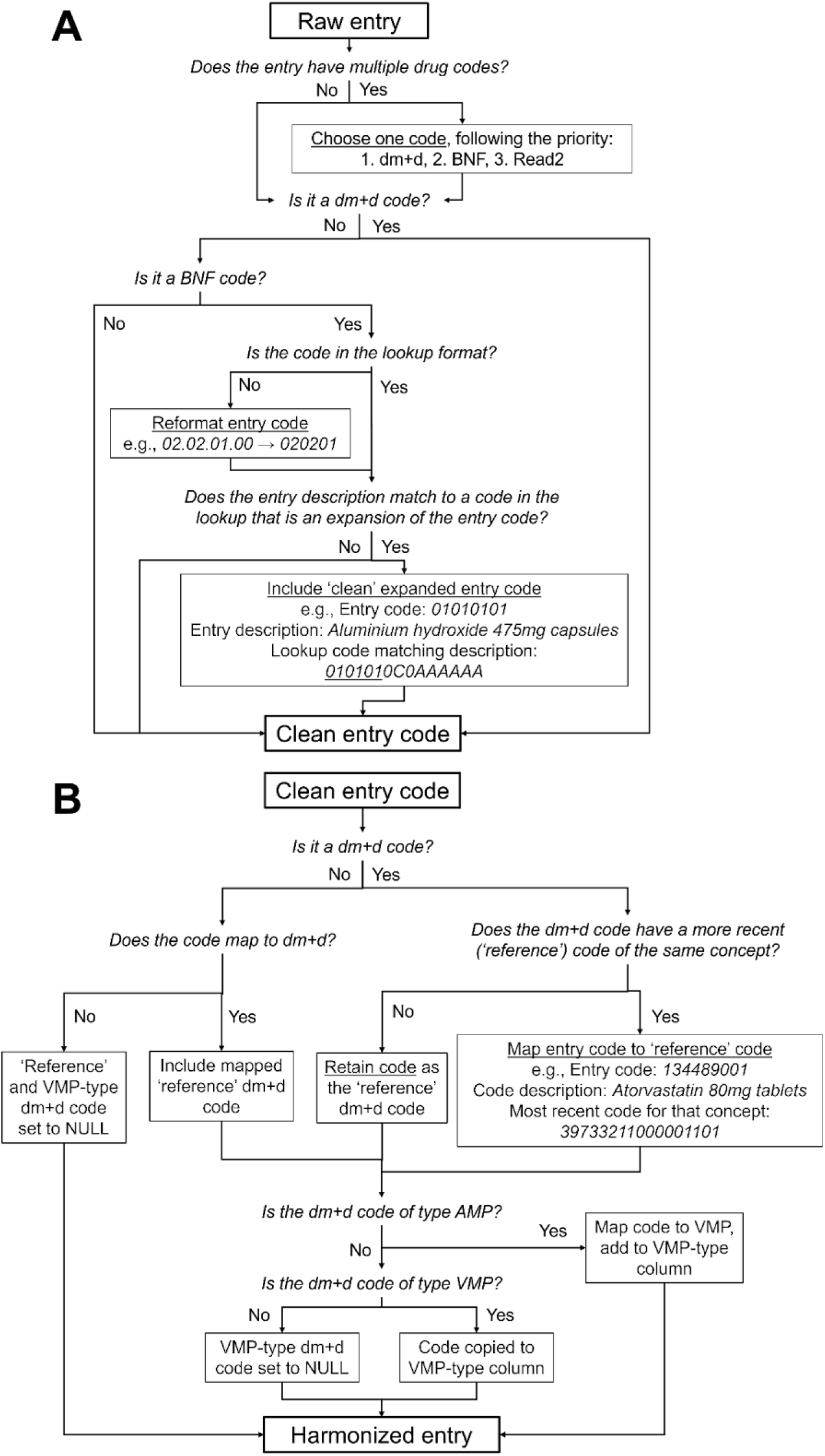
Preprocessing (A) and harmonisation (B) method flowcharts for UK Biobank primary care prescription records. By default, prescription codes not found in the lookup files will not have a mapped dm+d code. dm+d is the SNOMED CT dictionary of medicines and devices, BNF is the British National Formulary, Read2 is Read version 2, VMP is the dm+d Virtual Medicinal Product, AMP is the dm+d Actual Medicinal Product.

**Figure 2.**
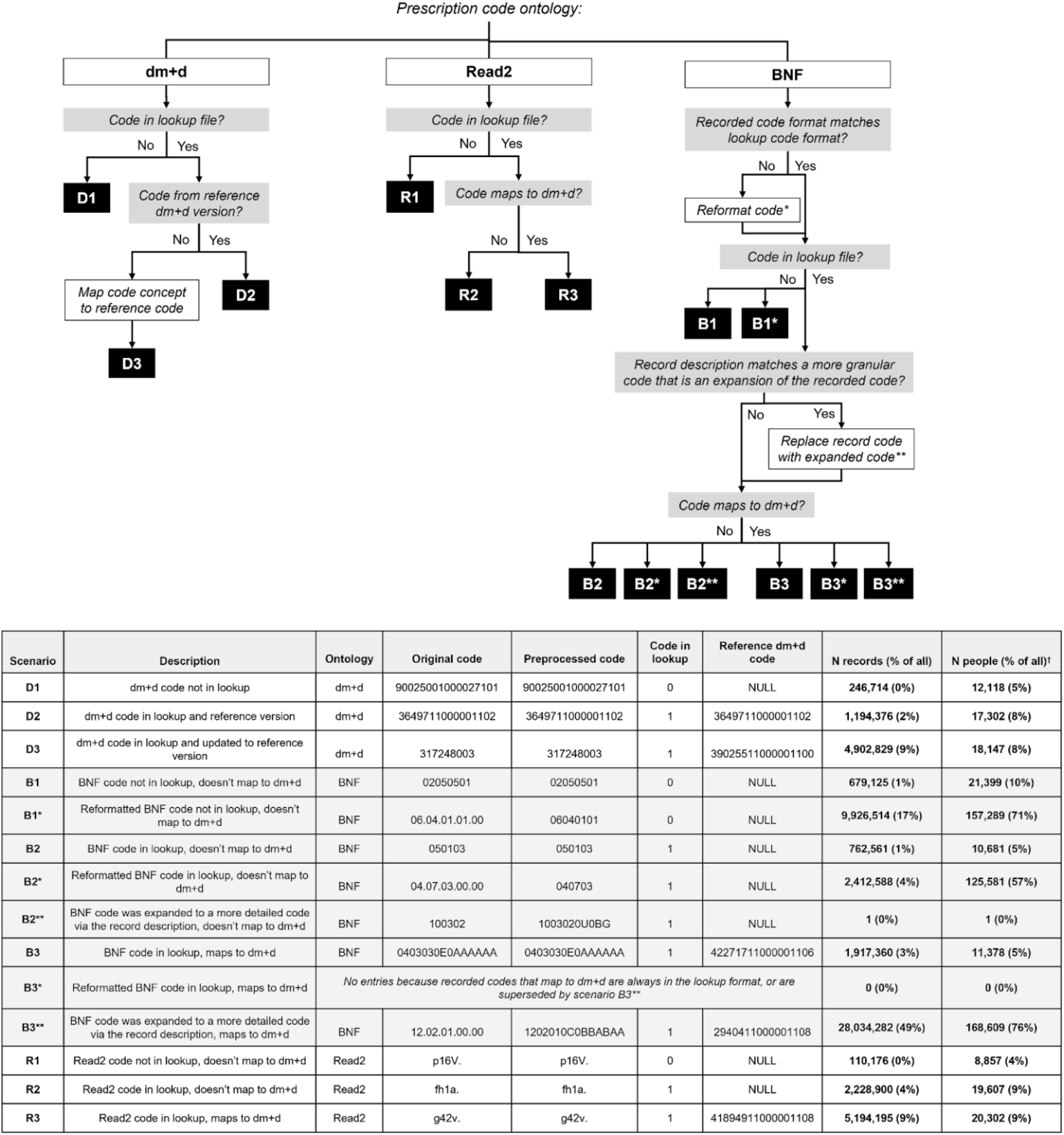
UK Biobank primary care prescription harmonisation scenarios per ontology of recorded drug. Reformatting (*) and expansion (**) examples shown in original versus preprocessed codes in inset table. Expansion supersedes formatting in scenario outcomes. dm+d is the SNOMED CT dictionary of medicines and devices, BNF is the British National Formulary, Read2 is Read version 2.

#### Preprocessing

We utilised the UKB prescription fields participant ID, data provider, issue date, and drug code(s) (read_2, bnf_code, dmd_code) from the “gp_prescriptions” table. We used drug name information to expand the granularity of BNF codes in certain scenarios (e.g., Figure 1A).

In cases where multiple ontologies were used to record a single prescription event, we implemented a preprocessing step to select the ontology with the highest priority based on a predefined order: 1. dm+d, 2. BNF, 3. Read2. The priority ontology was selected as dm+d because dm+d identifiers are codes from SNOMED CT, which is widely used worldwide and contains many well-defined relationships to other SNOMED CT concepts. It is also the NHS standard for medicines and medical devices in England, where the majority of the data originate. While BNF is useful due to its code structure containing innate information about drug components unlike dm+d, where each code is an integer with no inherent meaning, BNF is limited in interpretability due to UK-only scope. Read2 had the lowest priority because this system is obsolete in the NHS. Final drug codes were optionally preprocessed further to derive a ‘clean’ entry code; see details per ontology in the following harmonisation sections.

#### Read2 harmonisation

Read2 codes in English and Scottish entries were not considered because those entries either had BNF or dm+d codes that were preferentially selected to represent the drug concept. We could not locate a single authoritative reference standard for Welsh-based Read2 medication codes as these are locally-defined codes that are developed by primary care information system vendors. As a result, we developed a bespoke lookup file for Read2 medication codes to enable their mapping to dm+d. We queried the Unified Medical Language System (UMLS) [14] and filtered for these codes and their descriptions, as well as any SNOMED CT (dm+d) code that had the same UMLS concept identifier (CUI) as the Read2 code. For Welsh Read2 codes without a UMLS-based dm+d map, we compared the UMLS-based descriptions to reference dm+d descriptions and any exact matches were added to the mapping file. The mapped dm+d codes were finally aligned to their reference dm+d codes where necessary.

#### dm+d harmonisation

For prescription events recorded using dm+d where the dm+d concept used was inactive, we forward mapped the concepts to the most recent active concepts as defined in the authoritative lookup (see Supplemental Figure S2 and Table S2). The final stage of the process involved converting the mapped dm+d codes to a homogeneous granularity type, Virtual Medicinal Product (VMP). VMP codes are the most frequently used dm+d codes used in primary prescriptions and comprise of the medication information, strength and form. A subset of prescription events are recorded using AMP records which are based on VMP but represent the actual physical product (and include manufacturer information). To generate a dataset harmonised at the same level of information resolution we converted AMP codes to their VMP codes. After determining the reference dm+d code for all records, codes already in VMP-type were copied as the ‘reference VMP code’. AMP-type codes were mapped to their source VMP code using the dm+d lookups. Other types of dm+d codes (Virtual Therapeutic Moiety; <1% of records) were not mapped and had a NULL value as ‘reference VMP code’.

#### BNF harmonisation

For prescription events recorded in BNF, we followed two steps. Firstly, we reformatted BNF codes to a common format used in the lookup by removing period characters and any zeroes at the end of the code (i.e., broader codes where detailed information, like strength or chemical substance, is missing): for example, an entry code of ‘06.01.01.00’ was reformatted as ‘060101’. In some occasions the granularity of the code and the drug described in the description field were at odds. Often the drug description was identical to that of a code in the lookup that was an expansion of the event code. In these cases, we replaced the broad BNF with the more detailed code that matched the description and labelled as the ‘clean’ event code. Codes were not changed if the description was not exactly matched in the lookup, or if it matched to a BNF code that was not an expansion of the original code (ignoring the last character). For example, an entry with a code of ‘01010101’ and a description of ‘Aluminum hydroxide 475mg capsules’: the description matches the code ‘0101010C0AAAAAA’, and because the start of this code is the same as the entry code (ignoring the last character), the broad entry code (ignoring the last character, has a lookup description of ‘Antacids and simeticone’) is replaced by the more expanded code. Finally, we mapped BNF codes to dm+d where possible.

### validation and medication profiles

To validate the harmonised dataset and our overall methodology, we created and reviewed medication profiles that displayed contemporary medication prescribing in primary care for 312 physical and mental health diseases, selected to represent the disease burden and healthcare utilisation of the English population [15,16]. We used previously validated phenotyping algorithms [17] to identify disease cohorts in UKB using data from all available data sources (e.g., self reported, EHR, etc). For each disease, we matched disease cases to controls by age and sex (**Figure 3**). For cases and controls, we extracted and analyzed post-diagnosis harmonised prescription records. We removed records for drugs that were prescribed less than ten times or in fewer than five individuals. We compared medication frequency between case and control cohorts by quantifying the number of individuals prescribed each drug (represented as BNF subparagraph) in a 2×2 table.

**Figure 3.**
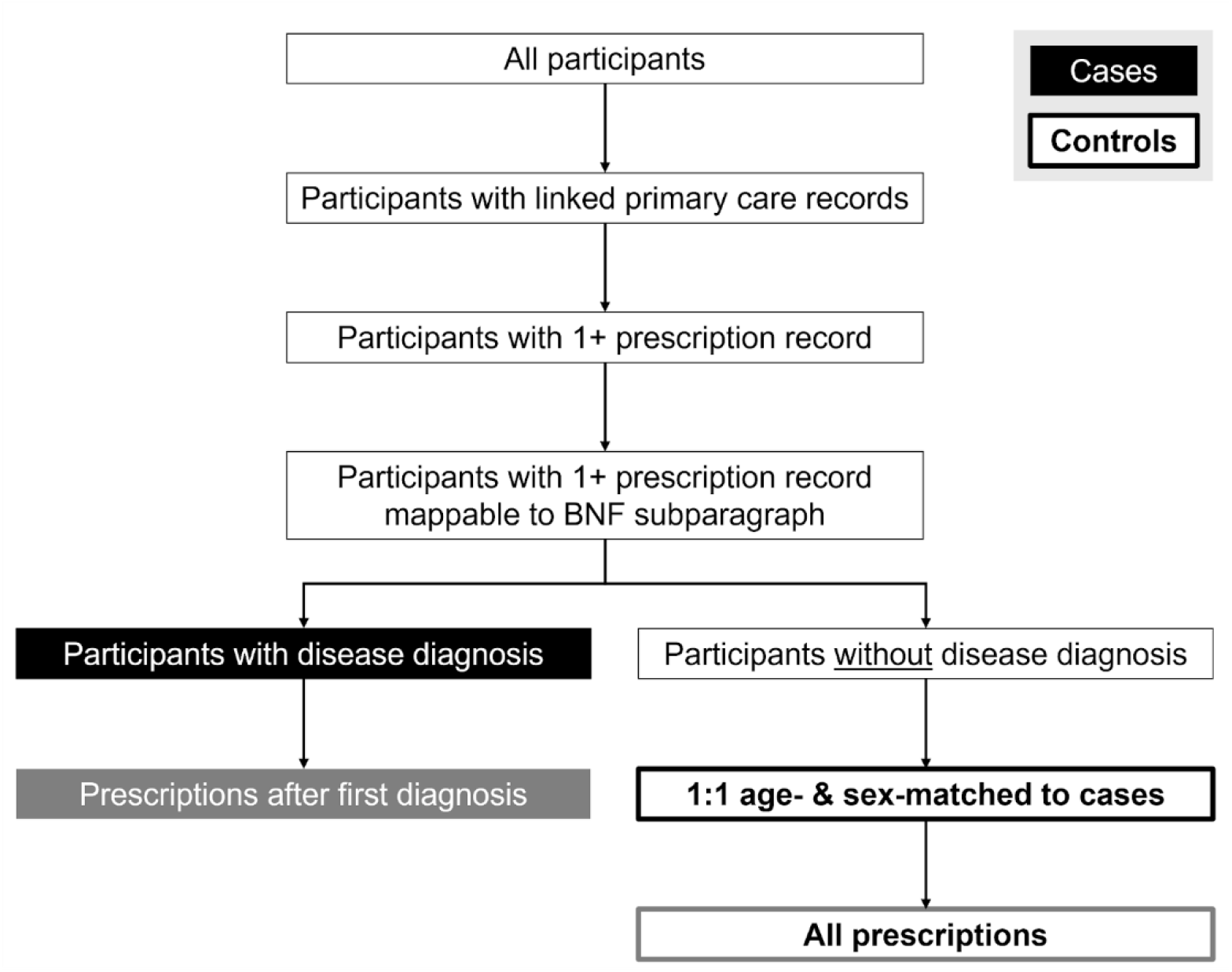
Flowchart of UK Biobank cohort analysis to determine medication profiles following prescription harmonisation. Controls contributed all available prescriptions rather than prescriptions after a matched index date as the primary purpose of this analysis is intended as a face-validity check of the harmonisation rather than a causal pharmacoepidemiological study. BNF is the British National Formulary.

Controls contributed all available prescriptions rather than prescriptions after a matched index date. This design choice was made because the medication profiles serve as a validation of the harmonisation methodology rather than as causal inference. The absence of an index date for controls means that control prescription histories are, if anything, more comprehensive than those of cases, which would attenuate disease-specific odds ratios and makes our approach conservative for detecting expected drug-indication signals.

We calculated and reported p-values and reported odds ratios (OR) using two-sided Fisher’s exact tests. We adjusted p-values for multiple comparisons using the Benjamini-Hochberg procedure [18] to control the false discovery rate (FDR) at 5%. We report the top ten BNF subparagraphs per disease, the strongest disease-medication associations, and the most commonly co-prescribed medications across all diseases.

### tools

Data were stored and managed using MySQL (v. 8.4). Analyses were performed using Python (v3.9.25).

## RESULTS

A Sankey diagram in **Figure 4** visualises the overall harmonisation procedure and breakdown of different factors/”datasets” indicated A-E: A) raw records from UK Biobank; B) preprocessed records; C) records in B where the drug code is in its ontology lookup file; D) records in C that have a reference dm+d code; E) records in D that map to/are VMP codes.

**Figure 4.**
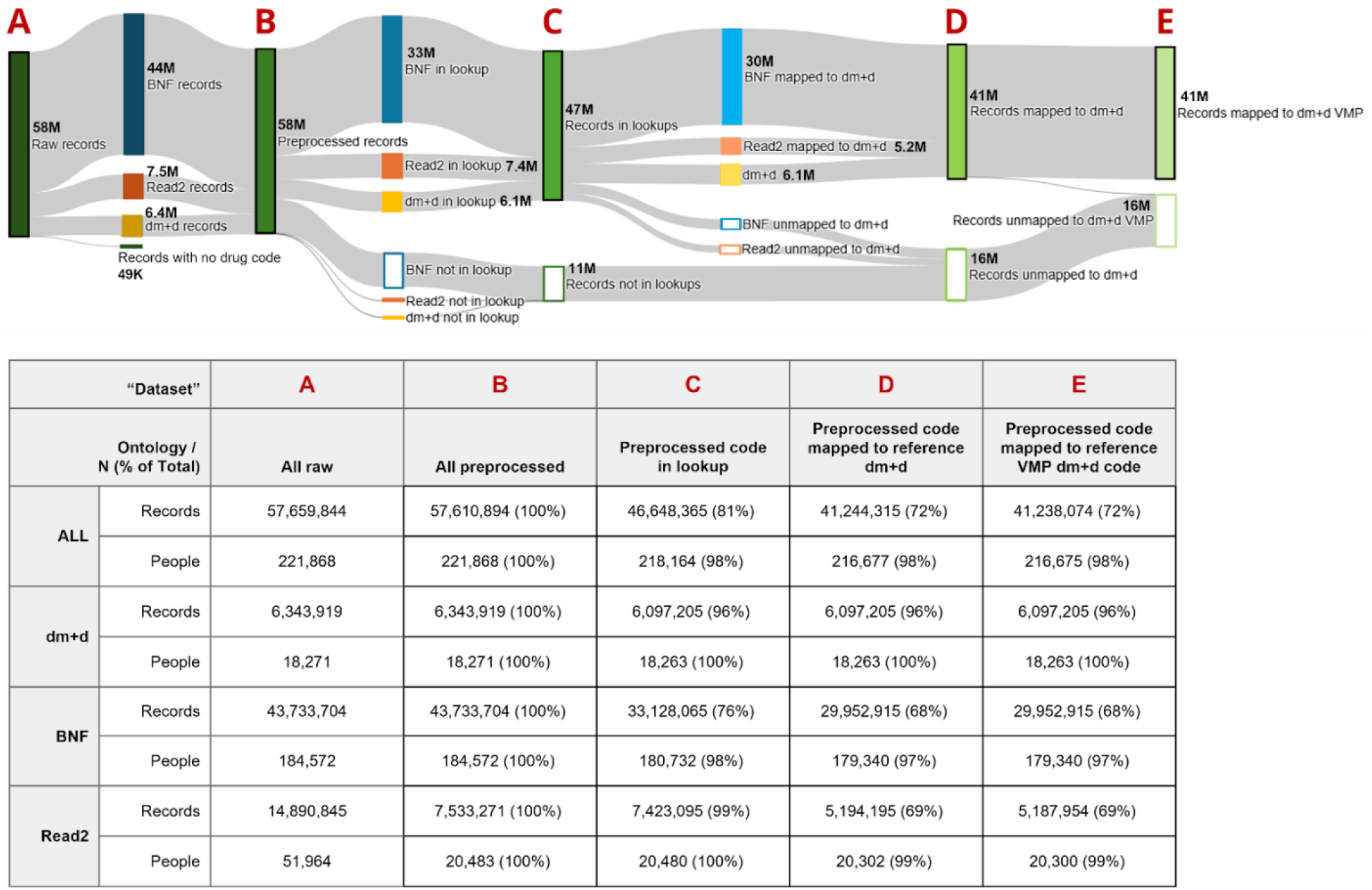
Sankey diagram of harmonisation procedure and breakdown of different factors/”datasets” indicated A-E: A) raw records from UK Biobank; B) preprocessed records; C) records in B where the drug code is in its ontology lookup file; D) records in C that have a reference dm+d code; E) records in D that map to/are VMP codes. Row-wise denominators in inset table for datasets B-E are the preprocessed (B) counts. Raw event counts and all people counts are non-exclusive. dm+d is the SNOMED CT dictionary of medicines and devices, BNF is the British National Formulary, Read2 is Read version 2, VMP is the dm+d Virtual Medicinal Product.

From the entire UKB cohort (N=502,131) we identified 221,868 (44%) individuals (55% female) with at least one primary care prescription record within a valid primary care registration period. We processed a total of 57,659,844 primary care prescription events: 6,343,919 recorded using dm+d, 43,733,704 recorded using BNF, and 14,890,845 recorded in Read2 codes (Table 1). The highest number of prescription events was contributed from the TPP provider in England (N=39,484,547).

Out of the total 57,659,844 events, the majority of raw prescription records (N=42,596,819; 74%) were coded using BNF alone. We identified 7,357,572 prescription records (13%) that used two different ontologies (Supplementary Figure S1). Specifically, we identified 6,220,687 records (11%) using both Read2 and dm+d and selected the dm+d codes for downstream mapping. Similarly, we identified 1,136,885 records (2%) that contained Read2 and BNF and prioritised BNF codes.

The application of the harmonisation algorithm yielded distinct scenarios across the three source ontologies that are detailed below with further examples in Figure 2. The most common harmonisation scenario (Scenario B3**) occurred when BNF codes were expanded to a more detailed code via the record description and then mapped to dm+d and was observed in 28,034,282 records (49%). For example, an entry with a BNF code of 12.02.01.00.00 and description of “Beconase Aqueous 50micrograms/dose nasal spray (GlaxoSmithKline UK Ltd)” was expanded to BNF concept 1202010C0BBABAA, which maps to dm+d code 2940411000001108. Records from England TPP data provider were processed using this scenario as we observed a mismatch in granularity between the total number of unique codes (n=635) and the total number of unique drug names (n=23,344).

For events recorded initially using dm+d, 246,714 records (0.004%) were absent from the lookup file (Scenario D1; e.g., undocumented code 90025001000027101), while 1,194,376 records (2%) were identified in the lookup and reference version (Scenario D2; e.g., 3649711000001102 “Colofac 50mg/5ml liquid”). A significant portion, 4,902,829 records (9%), required mapping codes to the reference version (Scenario D3). For example, the concept for “Ranitidine 75mg/5ml oral solution sugar free” was represented by 317248003 starting in 2014 (v1.0) but was replaced by 39025511000001100 in 2021 (v3.0) and the latter is therefore used as the reference code.

For records using Read2, 110,176 (0.002%) could not be found in the lookup (Scenario R1; e.g., undocumented code p16V.); however, 2,228,900 records (4%) were present but did not map to dm+d (Scenario R2; e.g., fh1a. “Nuvelle TS patch”), while 5,194,195 records (9%) successfully mapped to the target ontology (Scenario R3; e.g., g42v. “Clotrimazole 2% cream” maps to 41894911000001108).

We observed the highest potential number of harmonisation scenarios in BNF due to the formatting and expansion logic. Among BNF codes that did not map to dm+d, 679,125 (1%) failed primarily (Scenario B1; e.g., undocumented code 02050501), whereas a substantial 9,926,514 (17%) failed after reformatting (Scenario B1*; e.g., 06.04.01.01.00 reformatted to undocumented code 06040101). For BNF codes found in the lookup but not mapping to dm+d, we observed 762,561 records (1%) in the base format (Scenario B2; e.g., 050103 “Tetracyclines”), 2,412,588 records (4%) after reformatting (Scenario B2*; e.g., 04.07.03.00.00 reformatted to 040703 “Neuropathic pain”), and a single record (1.7E-8%) following expansion (Scenario B2**: 100302 expanded to 1003020U0BG “Voltarol Gel Patch”).

Finally, regarding successful mappings to dm+d from BNF, 1,917,360 records (3%) mapped directly (Scenario B3; e.g., 0403030E0AAAAAA “Fluoxetine 20mg capsules” maps to 42271711000001106) and zero records mapped via reformatting (Scenario B3*), while the expansion logic proved most critical, capturing the largest cohort of the study with 28,034,282 records (49%) successfully mapped (Scenario B3**; as described earlier).

The majority of individuals had at least one event code both referenced in the comprehensive lookups and mapped to dm+d (98%); records had 81% and 72% coverage, respectively. Only a very small number of records (N=48,950; 0.0008%) were completely dropped from the harmonised table; this was in cases where there was no drug code reported. Drug descriptions alone were not sufficient for harmonisation (e.g., **Supplementary Table S3**).

**Table 2** describes the breakdown of codes and records per data provider following harmonisation. A summary of the event and people counts for each of the described datasets is shown in Figure 4. Finally, **Supplementary Table S4** displays the records and people excluded from datasets B-E. People are only included in these counts if all of their prescription records met those specifications. **Supplementary Table S5** shows the breakdown of dm+d types of the reference codes in the dataset D. The last column shows the counts of homogeneous VMP records; effectively all of the information in the reference dm+d dataset can be converted to VMP.

**Table 2.**
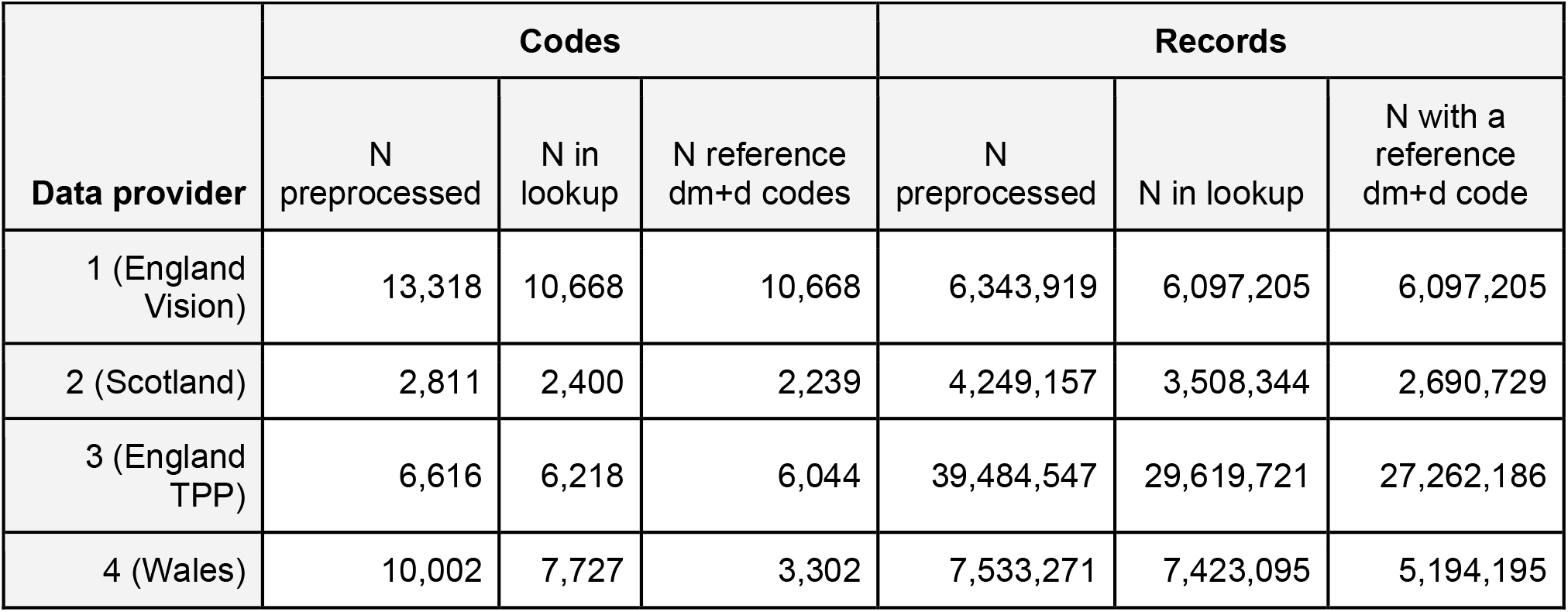
Breakdown of harmonised records per UK Biobank primary care prescription data provider. dm+d is the SNOMED CT dictionary of medicines and devices.

| Data provider | Codes |  |  | Records |  |  |
| --- | --- | --- | --- | --- | --- | --- |
|  | N preprocessed | N in lookup | N reference dm+d codes | N preprocessed | N in lookup | N with a reference dm+d code |
| 1 (England Vision) | 13,318 | 10,668 | 10,668 | 6,343,919 | 6,097,205 | 6,097,205 |
| 2 (Scotland) | 2,811 | 2,400 | 2,239 | 4,249,157 | 3,508,344 | 2,690,729 |
| 3 (England TPP) | 6,616 | 6,218 | 6,044 | 39,484,547 | 29,619,721 | 27,262,186 |
| 4 (Wales) | 10,002 | 7,727 | 3,302 | 7,533,271 | 7,423,095 | 5,194,195 |

Of the 13,780,789 BNF records that failed harmonisation to dm+d (24% of all records), 89% were from England TPP (n=12,222,361) and 11% were from Scotland (n=1,558,428), representing 31% and 37% of those providers’ records, respectively. These records failed harmonisation either because the encoded BNF drugs were too broad to be mapped to specific medications (e.g., 040703 ‘Neuropathic pain’) for 77% of unharmonised BNF records (n=10,605,639) or because the BNF code was not recognised in the lookup documentation (23%, n=3,175,150). For example, BNF code ‘02050501’ had 1,915,240 entries but is not listed in the BNF lookup file; these entries often had a more specific drug name entered (e.g., Lisinopril 5mg tablets), but because the drugs could not be verified against the unreferenceable and broad BNF code, they were not harmonised. Age and sex distributions of participants with BNF records that failed harmonisation to dm+d were consistent with the entire cohort: 45% male, 23% aged 40-49 at baseline, 33% aged 50-59, and 43% aged 60-69. Medication categories (BNF chapters) of these unharmonised records were similar to the distribution in all BNF entries: the cardiovascular system accounted for 33% of all BNF entries versus 28% of unharmonised BNF entries, the central nervous system had 15% versus 5% respectively, gastrointestinal had 9.3% versus 3.4%, endocrine had 11% versus 19%, musculoskeletal and joint diseases had 4.8% versus 3.2%, respiratory system had 6.3% versus 10%, skin had 4.2% versus 6.2%, infections had 3.8% versus 8.4%, and nutrition and blood had 4.0% versus 10%.

As mentioned, the Welsh Read2 records that failed harmonisation to dm+d were majority due to a missing map to dm+d (n=2,228,900; 95% of failed Read2 records). Welsh prescription records are comprised of local codes and do not contain drug descriptions, so drug and mapping information was limited to what was identifiable within the UMLS. All 20,483 participants with Welsh Read2 prescriptions had both harmonised and unharmonised drug records.

### validation and medication profiles

In 312 physical and mental health conditions, we identified 47657 associations with 239 unique drugs (represented as BNF subparagraphs). Of those, 23,352 associations (49.00%) survived statistical correction. The median OR of significant associations was 1.10 [IQR 0.53 to 2.01] and we observed an OR > 1 in 11,904 associations. Example results for a single disease are displayed for participants with chronic obstructive pulmonary disease (COPD), excluding bronchitis, in **Figure 5** with case and control cohorts summarised in **Table 3**.

**Table 3.** Summary of UK Biobank case and age- and sex-matched control cohorts for an example disease, COPD, following the procedure in Figure 3. Diagnostic code-based algorithm defined at Torralbo et al. (2025). *Age at first diagnosis.

|  | All UKB participants with COPD | COPD with primary care records | COPD with 1+ primary care prescription record AFTER first diagnosis | COPD with 1+ prescription record AFTER first diagnosis mappable to BNF subparagraph ('cases') |
| --- | --- | --- | --- | --- |
| N participants | 25,505 | 12,374 | 9,077 | 8,956 |
| N (%) male | 13,872 (54%) | 6,619 (53%) | 4,897 (54%) | 4,837 (54%) |
| N (%) female | 11,633 (46%) | 5,755 (47%) | 4,180 (46%) | 4,119 (46%) |
| N (%) age* <40 | 322 (1%) | 201 (2%) | 195 (2%) | 193 (2%) |
| N (%) age* 40-49 | 1,487 (6%) | 865 (7%) | 819 (9%) | 804 (9%) |
| N (%) age* 50-59 | 5,569 (22%) | 3,027 (24%) | 2,664 (29%) | 2,627 (29%) |
| N (%) age* 60-69 | 10,699 (42%) | 5,314 (43%) | 4,207 (46%) | 4,155 (46%) |
| N (%) age* 70-79 | 7,371 (29%) | 2,948 (24%) | 1,192 (13%) | 1,177 (13%) |
| N (%) age* 80+ | 185 (1%) | 66 (1%) | 0 (0%) | 0 (0%) |

**Figure 5.**
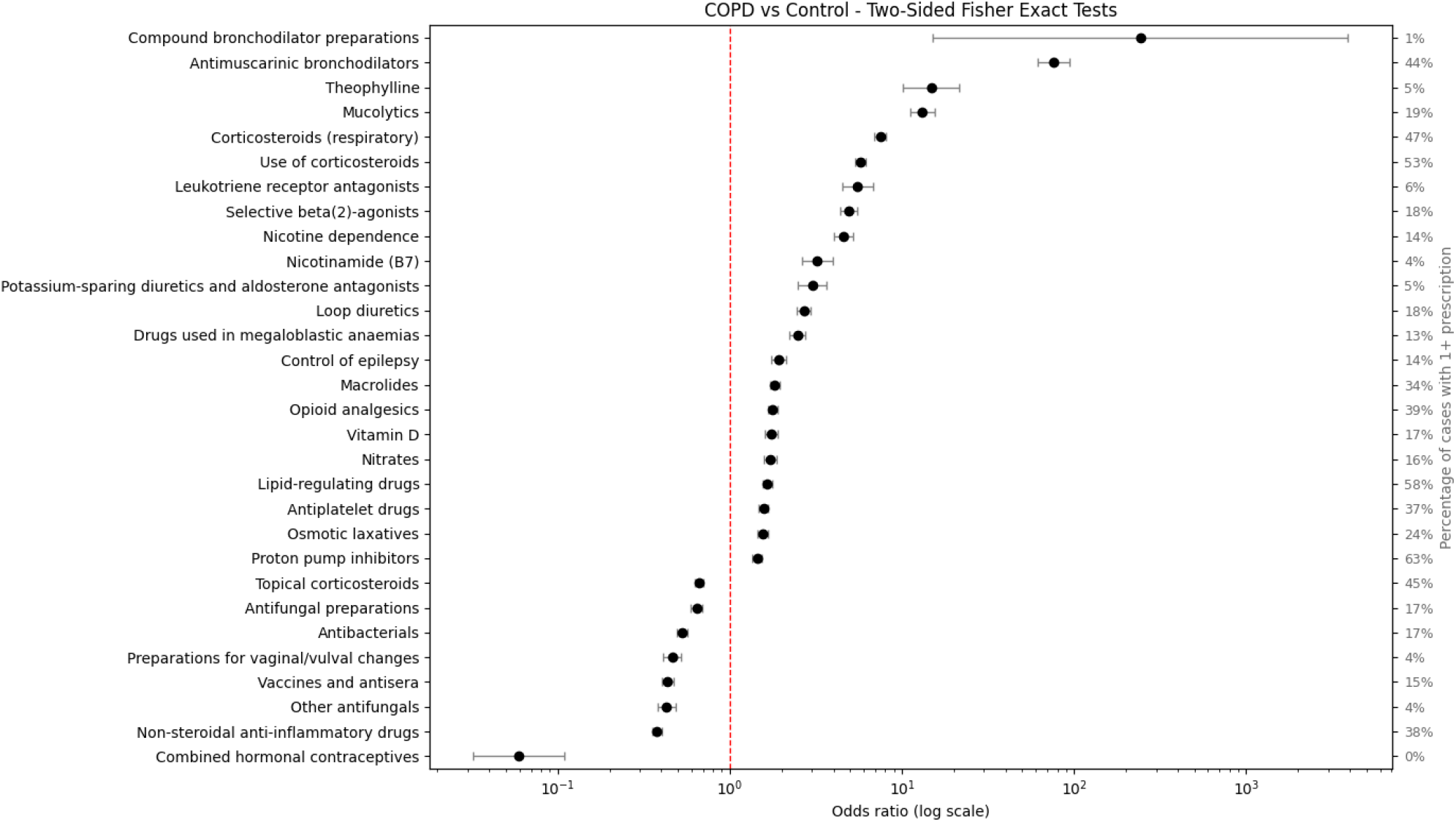
Odds ratios of drug categories (via BNF subparagraphs) being prescribed to cases versus controls, via two-sided Fisher’s exact tests, for example disease COPD. Two-sided Fisher’s exact tests were performed for 202 BNF subparagraphs, comparing 8,956 cases (individuals) against 8,956 controls (individuals). These tests analyzed a total of 2,560,769 case records and 1,911,835 control records. Results for 30 associations with lowest p-values shown.

The strongest associations between diseases and drugs were observed for Alzheimer’s disease and Drugs for dementia (OR 6081.7), Dementia and Drugs for Dementia (OR 3953.8), Parkinson’s disease and Dopaminergic drugs used in parkinsonism (OR 1100.4), Type 1 diabetes and Treatment of hypoglycaemia (OR 1049.1), and Myasthenia gravis with Drugs which enhance neuromuscular transmission (OR 782.6).

The most commonly associated medications shared across diseases were osmotic laxatives (n=231 diseases), vitamin D (n=211 diseases), stimulant laxatives (n=202 diseases), control of epilepsy medication (n=201) and drugs used in megaloblastic anaemias (e.g. folic acid) (n=199 diseases).

## DISCUSSION

Of the five types of dm+d codes, VTM and VMP, codes can change over time (e.g., Supplementary Table S2). Therefore the same drug concept can have multiple codes across different release versions and these may all be captured in the data due to the longitudinality of the records. VMP codes are the most common type of dm+d code used in primary care (Supplementary Table S1), so the issue of code versioning significantly contributed to fragmentation of dm+d records.

The large volume of BNF records that were mismatched in granularity between the recorded code and drug description (28 million) also highlight the value of our methodology, which judiciously expands the code granularity to match the described prescription when the broad code encapsulates the more specific named drug. Without this procedure, researchers run the risk of inappropriately matching drugs based on description alone, or omit detailed information if they solely use the broad coded values. This step greatly contributed to the value of the harmonised dataset and also enabled mapping to dm+d VMP concepts, which was impossible at the BNF level encoded in the raw records.

The medication profiles of diseases reflect contemporary clinical practice. In the top ten largest associations observed in the data across all diseases (**Supplementary Table 7**), we observed the highest enriched associations between Alzheimer’s and drugs for dementia (e.g., Galantamine), dementia and dementia medication (e.g., donepezil hydrochloride) and Parkinson’s disease and dopaminergic drugs used in parkinsonism (e.g., levodopa). Similarly, the analyses identified other cross-indicated therapies such as the use of hormone antagonists in primary prostate malignancies and the prescription of hypoglycaemia drugs as adjunctive therapy to insulin to improve glycemic control in individuals with type 1 diabetes.

Similarly, the medications observed for COPD (Figure 5) reflect current clinical practice in UK primary care, e.g., highest associations were observed for antimuscarinic bronchodilators (long-acting muscarinic antagonists like tiotropium) and theophylline which are the cornerstone of COPD maintenance therapy along with corticosteroids and long- and short-acting beta-2 agonists. The presence of nicotine dependence treatments reflects the management of smoking cessation and the use of macrolides indicates treatment of acute exacerbations of COPD. COPD has very high cardiovascular comorbidity which is evident by associations with loop diuretics, and vitamin D for heart failure and potassium-sparing diuretics, nitrates, lipid-regulating, and antiplatelet drugs for other CVD conditions. Nonsteroidal anti-inflammatory drugs (NSAIDs) can worsen bronchoconstrictions and are actively avoided in COPD patients which is illustrated by a negative association.

The most frequently co-occurring medications were those reflecting the systemic consequences of multimorbidity rather than disease-specific treatments. Laxatives, vitamin D supplementation, and drugs for megaloblastic anaemia ranked among the highest, consistent with the high prevalence of polypharmacy-induced constipation, nutritional deficiency secondary to chronic illness and corticosteroid use, and B12/folate depletion driven by metformin, methotrexate, and malabsorption syndromes. Antiepileptic drugs were 5th most commonly associated co-occurring medication reflecting their broad clinical utility across epilepsy, neuropathic pain, bipolar disorder, and anxiety which are all conditions highly prevalent in multimorbid populations.

While previous studies have utilised the rich self-reported prescription data offered by UKB, these have been primarily cross-sectional and to our knowledge none have curated a comprehensive drug-wide dataset using longitudinal records reflecting clinical care pathways. Previous studies have focused on certain disease- or intervention-specific prescriptions and manually filtered the dataset for matching records using methods like string matching of drug names [19], curating a list of codes [20,21], or a combination of the two [22]. The described methodology and results show that primary care prescribing data, as released by UK Biobank, can be significantly improved for research purposes by preprocessing and harmonising drug codes. This work also illustrates how the resulting ‘harmonised’ prescription dataset can be easily homogenised to a single level of information that enables agile research using the international ontology, dm+d (SNOMED CT). This dataset and its ease of filtering is far more comprehensive than common methods for filtering the raw records based on a manually-curated drug list, often only of one ontology or a list of drug names (for example, see multitude of values in raw records that were harmonised to a single drug concept in **Supplementary Table S8**).

This study has limitations. Many prescription records (3.1M, 6%) were recorded in the raw data using broad BNF levels that do not allow for harmonisation to dm+d concepts, or included codes that were unable to be referenced (11M, 19%). Missing information on local Welsh Read2 codes also led to 2.3M (4%) records unable to be harmonised. However, these records are retained in the harmonised dataset to allow for further study. It must also be noted that the source data describe only prescribed drugs, not dispensed medications, so findings from this data are not appropriate to use as determinations of what drugs people take. Diagnoses, clinical workflows, and prescription practices are likely to have changed over the study period but given that case ascertainment remains one of the primary use cases of UKB we have not performed temporal validation of the harmonization and have opted to process all data covering the study period. The approach to construct medication profiles potentially overestimates associations for commonly prescribed medications that accumulate with longer observation windows, and weakly positive or protective associations should be interpreted with this in mind.

Though this methodology is applied in UK-based ontologies (Read2, BNF) and data, the approach to harmonising electronic health records can be applied to other ontologies for mapping to SNOMED CT (e.g., expanding published lookup files, simplifying and expanding records based on available information). Converting invalid dm+d codes to a single ‘reference’ version and granularity (VMP) also enables streamlined analyses for any international supplier of dm+d prescription records. The harmonised dataset produced by this work is also more easily translated and directly comparable to other ontologies (e.g., Anatomical Therapeutic Code [ATC]) or other nations’ data where SNOMED CT prescriptions are available.

## CONCLUSIONS

Despite the value of prescription data for research, multiple problems are frequently encountered when using prescription records, including fragmentation (such as when multiple codes or levels of drug concepts are present) and data quality and completeness (such as incomplete reference files and undocumented or outdated codes); see also **Supplementary Table S6**. Our methodology addresses these problems by defragmenting and harmonising the available data via prioritising, expanding, mapping, and updating codes to a single reference version, thereby enabling population-wide analyses with minimal effort.

Harmonisation results show the increased value of conducting this procedure to both enrich the records supplied by UKB and also convert them to a single format that allows for the utilisation of the sophisticated SNOMED CT relationship system. Via the provided references to lookup and mapping files generated by this work, we hope that researchers who work with UK Biobank or other sources of primary care prescriptions can use our method to enable further applications of this resource for public good.

## Data Availability

We are unable to directly share source data from the UK Biobank due to their sensitive nature. Researchers can apply directly for access which is granted following protocol approval. To request access to UKB data, researchers need to apply directly to the UK Biobank, register in their access management system and submit a research study protocol https://www.ukbiobank.ac.uk/enable-your-research/apply-for-access.
All other analytical scripts and mapping files reported in this manuscript will be made available as open source upon publication.
Data from the medication profile analyses will be made publicly available upon publication.

## ETHICAL APPROVAL

UK Biobank data were obtained with approval of the UKB Access Management Committee protocol (https://bbams.ndph.ox.ac.uk/ams/) as part of Project 58356: Defining and redefining human disease at scale: an atlas of the human phenome.

## ACKNOWLEDGEMENTS

This research has been conducted using data from UK Biobank, a major biomedical database (www.ukbiobank.ac.uk). We thank the UK Biobank participants for making their health data available for research.

## DATA ACCESS

We are unable to directly share source data from the UK Biobank due to their sensitive nature. Researchers can apply directly for access which is granted following protocol approval. To request access to UKB data, researchers need to apply directly to the UK Biobank, register in their access management system and submit a research study protocol https://www.ukbiobank.ac.uk/enable-your-research/apply-for-access.

All other analytical scripts and mapping files reported in this manuscript have been made available as open source in the following repository: To be shared upon publication

Data from the medication profile analyses is publicly available in the following repository: To be shared upon publication

