## Supplementary Material for "Harmonising UK primary care prescription records for research: A case study in the UK Biobank"

### Supplementary Figures

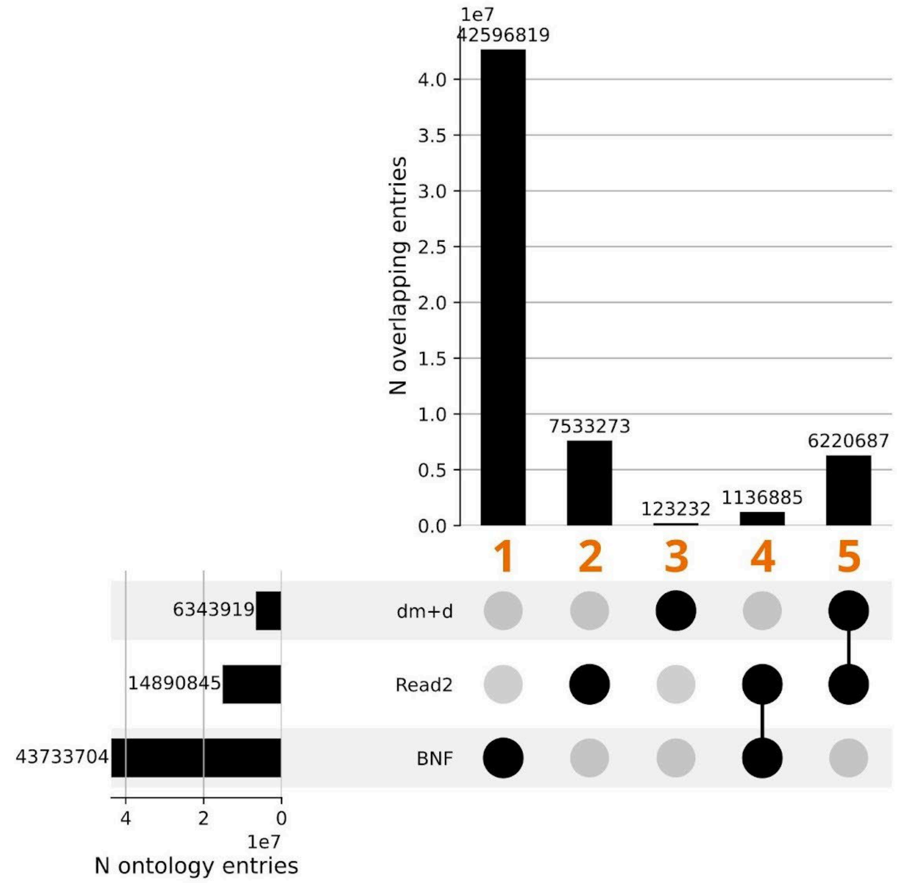


| Entry group | Harmonized ontology |
|---|---|
| 1 | BNF |
| 2 | Read2 |
| 3 | dm+d |
| 4 | BNF |
| 5 | dm+d |

**Figure S1**. Ontologies present in the raw UK Biobank primary care prescription records. In cases where multiple ontologies were present, a single code for the event was chosen following the logic in the inset table. dm+d is the SNOMED CT dictionary of medicines and devices, BNF is the British National Formulary, Read2 is Read version 2.

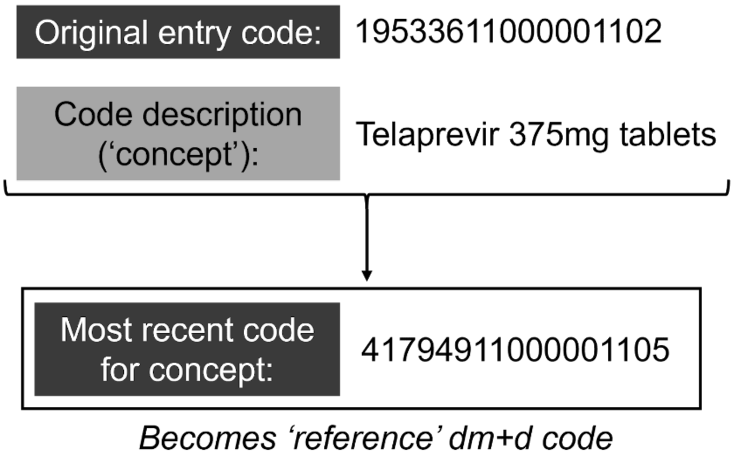


**Figure S2.** Example of selecting a reference dm+d code to reflect a concept in an entry with an outdated code. dm+d is the SNOMED CT dictionary of medicines and devices.

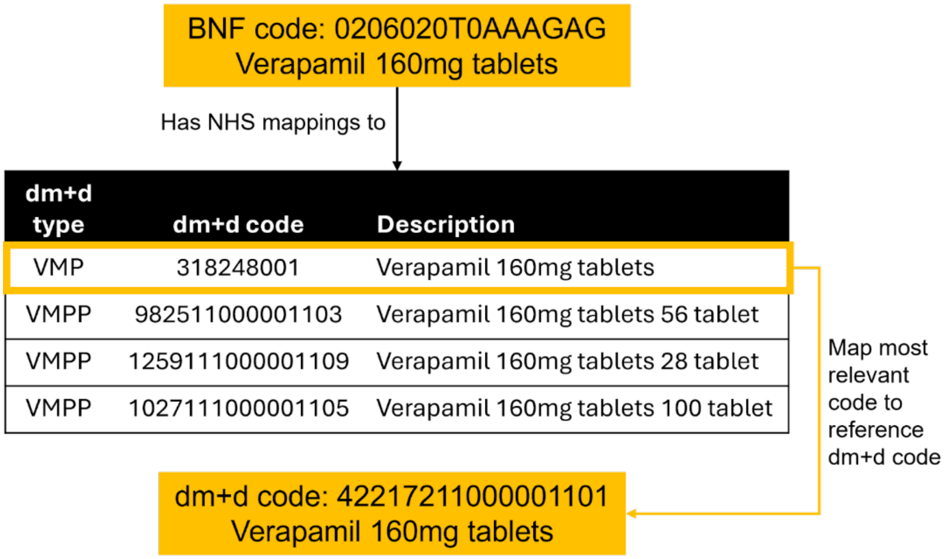


**Figure S3**. Example of BNF to dm+d code mapping. dm+d is the SNOMED CT dictionary of medicines and devices, BNF is the British National Formulary, VMP is Virtual Medicinal Product, VMPP is Virtual Medicinal Product Pack.

### Supplementary Tables

**Table S1.** Examples of dm+d prescription code types and their occurrence in the raw UKB data. The code types are further described in the dm+d data model (NHS Business Services Authority, 2024).

| dm+d code type | Abbreviation | Example code description | N UKB records | N UKB codes |
|---|---|---|---|---|
| Virtual Therapeutic Moiety | VTM | Amoxicillin | 0 | 0 |
| Virtual Medicinal Product | VMP | Amoxicillin 500mg capsules | 5,070,319 | 3,694 |
| Virtual Medicinal Product Pack | VMPP | Amoxicillin 500mg capsules 21 capsule | 0 | 0 |
| Actual Medicinal Product | AMP | Amoxicillin 500mg capsules (GlaxoSmithKline) | 1,026,886 | 6,974 |
| Actual Medicinal Product Pack | AMPP | Amoxicillin 500mg capsules (GlaxoSmithKline) 21 capsule | 0 | 0 |

**Table S2.** Example of logic for selecting 'reference' dm+d codes using the authoritative dm+d lookup. The most recent code for the concept is used as the reference.

| dm+d code | Description | Valid years (versions) | Previous dm+d code | Reference dm+d code |
|---|---|---|---|---|
| 19533611000001102 | Telaprevir 375mg tablets | 2014 (1.0) - 2015 (4.0) | N/A | 41794911000001105 |
| 704467005 | Telaprevir 375mg tablets | 2015 (4.1) - 2023 (5.2) | 19533611000001102 | 41794911000001105 |

| dm+d code | Description | Valid years (versions) | Previous dm+d code | Reference dm+d code |
|---|---|---|---|---|
| 41794911000001105 | Telaprevir 375mg tablets | 2023 (10.0) - present (2024, 7.2) | 704467005 | 41794911000001105 |

**Table S3.** Examples of raw UK Biobank primary care prescription entries that were not harmonised due to lack of drug code.

| read_2 | bnf_code | dmd_code | drug_name |
|---|---|---|---|
| NULL | NULL | NULL | Triptorelin Acetate Powder and solvent for injection 4.2 mg vial (3 mg Triptorelin) |
| NULL | NULL | NULL | Liquid Paraffin And White Soft Paraffin Ointment 50 % + 50 % |
| NULL | NULL | NULL | Elasticated Tubular Bandage Bp 10 cm size F |

**Table S4.** Summary of UK Biobank primary care prescription records excluded following harmonisation. Percentage denominator is ‘all’ counts per labelled dataset (see Figure 4). dm+d is the SNOMED CT dictionary of medicines and devices, VMP is the Virtual Medicinal Product form of dm+d codes.

| “Dataset” | B | C | D | E |
|---|---|---|---|---|
| **N (% of total dataset)** | **Record has no drug code** | **Code not in lookup** | **Code unmapped to dm+d** | **Code unmapped to dm+d VMP** |
| Records | 48,970 (0%) | 10,962,529 (19%) | 16,366,579 (28%) | 16,372,820 (28%) |
| People | 0 (0%) | 3,704 (2%) | 5,191 (2%) | 5,193 (2%) |

**Table S5.** Summary of reference dm+d codes in harmonised UK Biobank primary care prescription records by dm+d type. *Denominator is all records with a reference dm+d code (dataset D in Figure 4). VTM is Virtual Therapeutic Moiety, VMP is Virtual Medicinal Product, and AMP is Actual Medicinal Product.

| | VTM | VMP | AMP |
|---|---|---|---|
| N People (% of all*) | 1,127 (1%) | 214,138 (97%) | 181,178 (82%) |
| N Records (% of all*) | 6,241 (0%) | 36,324,022 (88%) | 4,914,052 (12%) |
| N Codes (%) | 76 (0.5%) | 4,620 (35%) | 8,484 (64%) |

**Table S6.** Problems and this study’s solutions to using primary care prescriptions for research. dm+d is the SNOMED CT dictionary of medicines and devices, BNF is the British National Formulary, Read2 is Read version 2.

| Problem | Example | Solution | Example |
|---|---|---|---|
| **Fragmentation** | Multiple drug codes from different ontologies within a single event | Preprocessing | Single prioritised code per event (1. dm+d, 2. BNF, 3. Read2) |
| | | Harmonisation | Preprocessed codes mapped to the ‘reference’ dm+d version where possible |

| Problem | Example | Solution | Example |
|---|---|---|---|
| | Equivalent concepts encoded at different levels | Homogeneous dataset | All possible records mapped to dm+d concepts at the Virtual Medicinal Product level of drug information. |
| **Data quality and completeness** | Missing or incomplete reference files | This study | Novel, open-source lookup files generated by this process:<br>• consolidated dm+d lookup & map to single version ('reference') of concept codes<br>• expanded BNF to dm+d map<br>• Welsh Read2 lookup file<br>• Welsh Read2 to dm+d map |
| | Recorded codes cannot be referenced or contain conflicting information | Preprocessing | BNF codes consistently formatted to match lookup |
| | | | Expanded BNF code granularity where event code is abbreviated form of the lookup code matching the event description |
| | | | Simple flag for whether the event code is referenceable in its ontology lookup file |
| | Outdated codes | Harmonisation | dm+d codes mapped to same concept coded in a single 'reference' version |

**Table S7.** Ten highest disease-drug associations observed in UK Biobank primary care prescriptions of participants in various disease cohorts.

| Disease | BNF Subparagraph | Drug name | Odds ratio (OR) | P-value (adjusted) |
|---|---|---|---|---|
| Alzheimer's | 411000 | Drugs for dementia | 6081.8 | 7.20E-289 |
| Dementia | 411000 | Drugs for dementia | 3953.8 | 0.00E-02 |
| Parkinson's | 409010 | Dopaminergic drugs used in parkinsonism | 1100.4 | 0 |
| Diabetes Type I | 601040 | Treatment of hypoglycaemia | 1049.9 | 1.23E-132 |
| Myasthenia | 1002010 | Drugs which enhance neuromuscular transmission | 782.7 | 1.32E-48 |
| Autoimmune liver disease | 109010 | Drugs affecting biliary composition and flow | 778.0 | 2.94E-71 |
| Pancreatic cancer | 109040 | Pancreatin | 745.6 | 3.82E-62 |
| Diabetes Type II | 601021 | Sulfonylureas | 732.1 | 0 |
| Diabetes (any) | 601011 | Short-acting insulins | 724.4 | 9.06E-106 |
| Schizophrenia | 409020 | Antimuscarinic drugs used in parkinsonism | 689.6 | 1.31E-83 |

**Table S8.** Contents of raw UK Biobank primary care prescription records ("gp_prescriptions") matched to 'Paracetamol 500mg tablets' via the harmonised table.

| Field | N unique values | Examples |
|---|---|---|
| read_2 | 2 | di21.00; di21. |
| bnf_code | 2 | 04.07.01.02.00; 04070100 |
| dmd_code | 13 | 322236009; 171811000001104; 13429711000001102; 67111000001103; … |
| drug_name | 38 | PARACETAMOL tabs 500mg; Paracetamol 500mg caplets (A A H Pharmaceuticals Ltd); Panadol Advance 500mg tablets (GlaxoSmithKline Consumer Healthcare); … |
